# Automating Screening of Titles and Abstracts in Systematic Reviews: An Assessment of GPT-4o mini

**DOI:** 10.64898/2026.05.15.26353334

**Authors:** Mir Sohail Fazeli, Ellen Kasireddy, Mir-Masoud Pourrahmat, Cuthbert Chow, Jean-Paul Collet

## Abstract

**Background:** Systematic literature reviews (SLRs) are essential in medical research, but are often time-consuming and costly, necessitating more efficient methods while maintaining accuracy.

**Objective:** This study assessed the performance of a GPT-4o mini large language model (LLM) in automating the first phase of study selection based on titles and abstracts in systematic reviews. Specifically, we evaluated whether the model improved efficiency without compromising on quality.

**Methods:** Structured prompts were created for a GPT-4o mini LLM to facilitate title and abstract screening. The model’s performance was evaluated against expert human reviewers across five systematic reviews on inclusion rates, sensitivity, specificity, accuracy, positive predictive value, and negative predictive value.

**Results:** The model screened a total of 15,605 records. It included a higher percentage of studies than human screeners, with 3.5% (n=549/15,605) true positives and 14.2% (n=2,218/15,605) false positives. The model achieved an overall accuracy of 85.1%, with a sensitivity of 83.2% and specificity of 85.2%. The positive predictive value was 19.8%, while the negative predictive value was 99.1%. The model was able to screen 1,000 titles and abstracts in 40 minutes, compared to 16 hours required by a human reviewer.

**Conclusion:** This study demonstrated a strong performance and efficiency in the automation of title and abstract screening in SLRs using an advanced LLM. Further refinements could optimize the balance between sensitivity and specificity, supporting broader implementation in evidence synthesis. A hybrid AI-human approach is recommended to ensure accuracy, reduce reviewer burden, and maintain the methodological rigor required for high-quality SLRs.

## Introduction

Systematic literature reviews (SLR) in medical research play a crucial role in compiling existing evidence to guide clinical practice and policymaking. They provide a meticulous synthesis of study findings, enabling clinicians to make well-informed, evidence-based decisions while identifying gaps in current knowledge.[1, 2] However, conducting systematic reviews can be highly labor-intensive and costly. The process involves extensive literature searches and screening of identified records, critical evaluation of publications, and precise data extraction, all requiring significant expertise and effort.[3, 4] Consequently, this can lead to delays in disseminating findings and challenges in maintaining up-to-date information.

Recent advancements in artificial intelligence (AI), particularly large language models (LLMs) like OpenAI’s GPT-4 series, offer the potential to streamline aspects of the systematic review process. These AI-driven tools can help refine search strategies, identify relevant publications, and filter information, ultimately reducing completion times while minimizing error.[5–9] By enabling researchers to manage increasingly complex datasets, LLMs help maintain the relevance and comprehensiveness of systematic reviews in fast-evolving fields. As AI tools continue to advance, their integration into the systematic review process is expected to be more efficient, cost-effective, and accessible to a broader research community, ultimately supporting more informed decision-making across various disciplines.[10, 11]

Despite these promising advances, the application of AI tools in systematic reviews remains at an early stage and raises important concerns. Chief among them is the question of whether these models can reliably identify all relevant studies without introducing systematic errors or omissions. While LLMs can dramatically increase screening speed, it remains unclear whether this efficiency compromises the comprehensiveness and accuracy of the selection process, particularly in identifying nuanced or borderline cases that require human domain expertise. Given the high stakes of evidence synthesis in health and medical research, it is essential to rigorously evaluate the performance of these AI models before integrating them into standard review workflows.

This study explores the integration of GPT-4o mini for screening articles during the first phase of study selection in systematic reviews based on article titles and abstracts, aiming to evaluate its effectiveness in improving efficiency while upholding the quality of study selection. By leveraging AI, we seek to demonstrate a practical application that has the potential to transform traditional methodologies, making the systematic review process more efficient and reliable.

## Methods

### Model Development

A team of scientists and experts in SLR methodologies developed a customized LLM-based model powered by the GPT-4o mini engine. The model was designed to perform the role of a research scientist, specifically to support title and abstract screening in accordance with predefined SLR objectives and Population, Intervention, Comparison, and Outcome (PICO) criteria.

A structured prompt engineering approach was used to adapt the model for title and abstract screening. The model was iteratively tested on a set of ∼100-150 title and abstract records drawn from previously conducted SLRs. These records were screened by experienced reviewers and were used exclusively to guide prompt development. Iteration continued until the model’s decisions consistently matched expert expectations across a test set of records.

Prompts were refined over multiple rounds to improve performance, focusing on decision consistency and clarity of justification. Final prompt structures (as provided in **Supplementary Table S1**) ensured that the model provided a binary inclusion or exclusion decision for each study, along with a justification and, where applicable, reasons for exclusion mapped to the relevant PICO element.

### Model Configuration

To manage context window limits and maintain accuracy, screening was conducted in batches of 10 records. The model was run with a temperature setting of 0, which reduces randomness in the model’s responses and leads to more deterministic, consistent outputs. The model was run with the highest level of strictness, which refers to the model’s threshold for excluding studies based on the PICO criteria: higher strictness levels result in more conservative inclusion behavior (i.e., greater likelihood of excluding records).

### Evaluation Dataset

The model was evaluated using five published systematic reviews across diverse therapeutic areas (See **Supplementary Table S2-S6** for PICO tables): two reviews on humanistic burden,[12, 13] two reviews on clinical burden,[14, 15] and one review on health state utility values.[16] These reviews covered a range of therapeutic areas, including oncology (two reviews), type 1 diabetes, schizophrenia, and mobility impairment (one review each). Four[12–15] of the five reviews did not allow exclusion based on outcomes, whereas one review[16] did. The model was provided with the same inclusion and exclusion criteria as each review, including whether outcome-based exclusions were permitted.

Each review followed Cochrane guidelines, which recommend a two-stage study selection process involving independent screening by two reviewers. Generally, in the first stage of study selection, titles and abstracts are screened to exclude clearly irrelevant studies based on predefined eligibility criteria aligned with the PICO framework. Discrepancies between reviewers are resolved through discussion to reach consensus.[17] Although full-text screening is a subsequent stage to confirm study eligibility, the present evaluation focuses solely on the title and abstract screening stage.

### Model Evaluation Metrics

In the context of using the model for title and abstract screening in a systematic review, key classification metrics were defined as follows:

- True Positives: Cases where the model correctly identified and included a publication that human reviewers had also included.
- False Positives: Cases where the model mistakenly included a publication that human reviewers had excluded.

- Together, true positives and false positives represent the model inclusion rate, reflecting the percentage of publications deemed “Included” based on the specified PICO criteria.
- True Negatives: Cases where the model correctly identified and excluded a publication that human reviewers had also excluded.
- False Negatives: Cases where the model mistakenly excluded a publication that human reviewers had included.

- Together, true negatives and false negatives represent the model exclusion rate, reflecting the percentage of publications deemed “Excluded” based on the specified PICO criteria.

The following metrics were used to assess the model’s performance relative to human reviewers:

- Sensitivity (Recall or True Positive Rate): The model’s ability to correctly identify relevant publications. The formula used for this calculation was as follows:

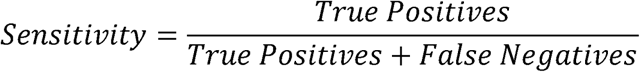

- Specificity (True Negative Rate): The model’s ability to correctly identify irrelevant publications. The formula used for this calculation was as follows:

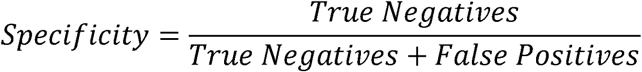

- Positive Predictive Value (PPV): The proportion of publications included by the AI model that were truly relevant. The formula used for this calculation was as follows:

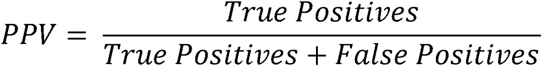

- Negative Predictive Value (NPV): The proportion of publications excluded by the AI model that were truly irrelevant. The formula for this calculation was as follows:

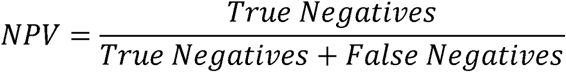

- Accuracy: Measured the proportion of correctly classified records (both included and excluded) out of the total number of records screened, calculated as follows:

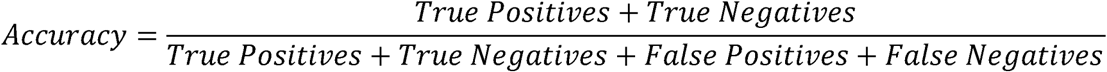

Classification and performance metrics were assessed at two levels: overall (by summing true positives, false positives, true negatives, and false negatives across all five reviews) and individually for each review.

### Efficiency Assessment

In addition to the above performance metrics, efficiency was assessed by measuring the time taken to screen 1,000 titles and abstracts using each method. The comparison included screening by a single human reviewer and by the model. Time savings were calculated as the difference in total screening time between the model and the human reviewer.

## Results

### Model Output

The model screened 15,605 title and abstract records across the five systematic reviews. The largest number of records came from the review on clinical burden of mobility impairment (n=6,156),[15] followed by reviews on the humanistic burden in patients with schizophrenia (n=3,356),[13] clinical burden of renal carcinoma (n=2,970),[14] humanistic burden in children with type 1 diabetes (n=1837),[12] and health state utility values of cancer stages (n=1,286)[16] (**Table 1**).

**Table 1:** Review-level performance metrics of the model for title/abstract screening.

| Review | Review description | Total number of abstracts screened | TP | FP | TN | FN | Sensitivity | Specificity | Accuracy | PPV | NPV |
| --- | --- | --- | --- | --- | --- | --- | --- | --- | --- | --- | --- |
| Allen et al. 2022[12] | Humanistic burden in children with type 1 diabetes | 1,837 | 35 (1.9%) | 448 (24.4%) | 1,353 (73.7%) | 1 (0.1%) | 97.2% | 75.1% | 75.6% | 7.2% | 99.9% |
| Bruhn et al. 2022[13] | Humanistic burden in patients with schizophrenia | 3,356 | 43 (1.3%) | 850 (25.3%) | 2,455 (73.2%) | 8 (0.2%) | 84.3% | 74.3% | 74.4% | 4.8% | 99.7% |
| Ejzykowicz et al. 2022[14] | Clinical burden of renal cell carcinoma | 2,970 | 56 (1.9%) | 47 (1.6%) | 2,864 (96.4%) | 3 (0.1%) | 94.9% | 98.4% | 98.3% | 54.4% | 99.9% |
| Guo et al. 2022[15] | Clinical burden of mobility impairment | 6,156 | 288 (4.7%) | 650 (10.6%) | 5,128 (83.3%) | 90 (1.5%) | 76.2% | 88.8% | 88.0% | 30.7% | 98.3% |
| Pourrahmat et al. 2021[16] | Health state utility values of cancer stages | 1,286 | 127 (9.9%) | 223 (17.3%) | 927 (72.1%) | 9 (0.7%) | 93.4% | 80.6% | 82.0% | 36.3% | 99.0% |
| Overall | N/A | 15,605 | 549 (3.5%) | 2,218 (14.2%) | 12,727 (81.6%) | 111 (0.7%) | 83.2% | 85.2% | 85.1% | 19.8% | 99.1% |

FN, number of false negatives; FP, number of false positives; N/A, not applicable. NPV, negative predictive value; PPV, positive predictive value; TN, number of true negatives; TP, number of true positives.

### Proportion of Included Publications

Across the five systematic reviews, the model included 17.7% (n=2,767/15,605) of records, with the individual inclusion rate ranging from 3.5%[14] to 27.2%.[16] The model inclusion rate was higher than that of human reviewers for all reviews, with 3.5% (n=549/15,605) true positives and 14.2% (n=2,218/15,605) false positives (**Fig 1**; **Table 1**).

**Fig 1:**
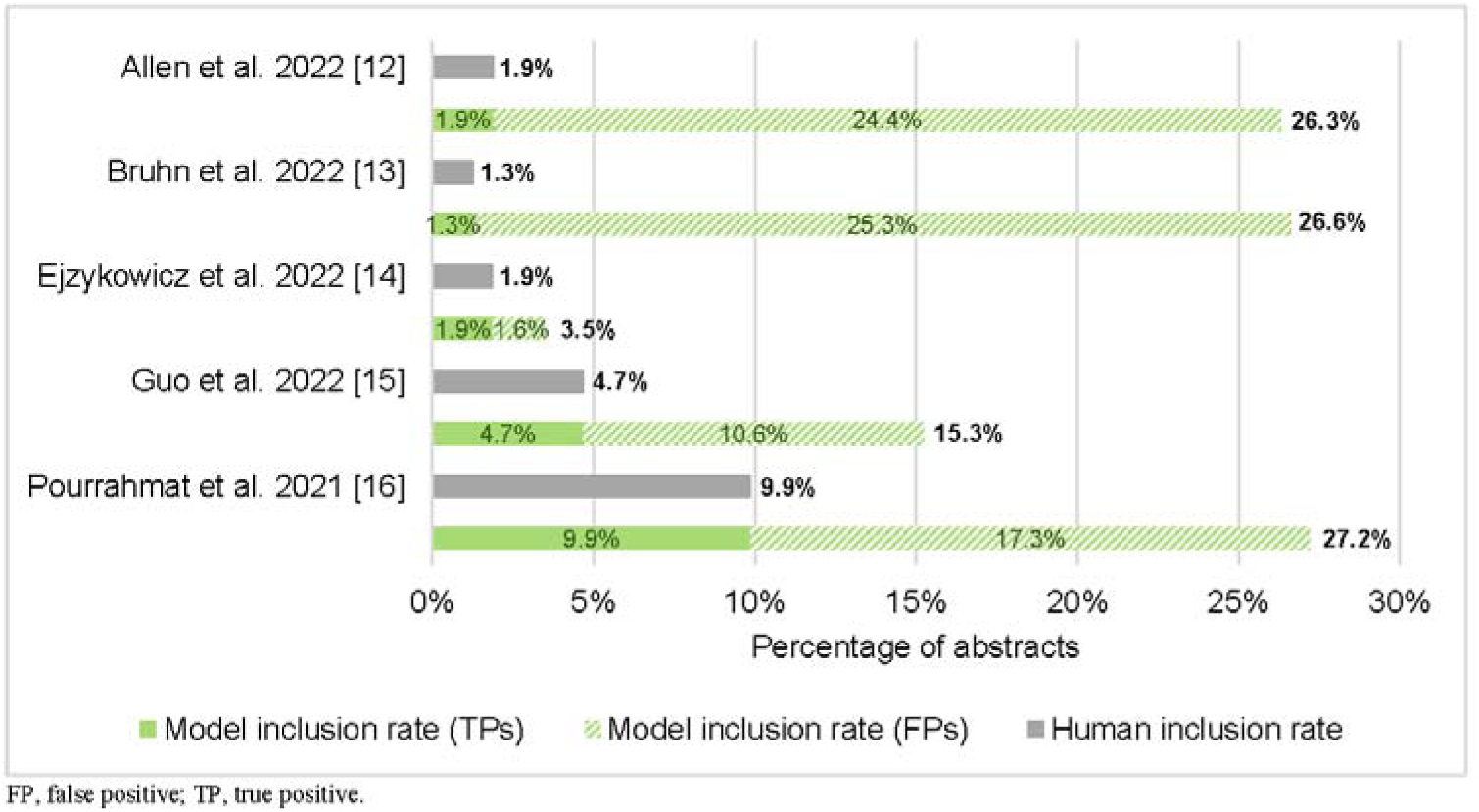
Comparison of review-level human and model inclusion rates from title and abstract screening

Notably, the model inclusion rate was substantially higher than manual screening for the two systematic reviews on humanistic burden. In the review by Allen et al. [12] (humanistic burden in children with type 1 diabetes), the AI-assisted screening resulted in an inclusion rate of 26.3%, compared to 1.9% with manual screening. Similarly, in the review by Bruhn et al. [13] (humanistic burden in patients with schizophrenia), the model reported an inclusion rate of 26.6%, whereas human reviewers included only 1.3% of records.

### Exclusion Rate and Reasons for Exclusions

Across the five systematic reviews, the model excluded 82.3% (n=12,838/15,605) of records, with the individual exclusion rate ranging from 72.8%[16] to 96.5%.[14] Among the excluded records, the true exclusion rate (i.e., NPV) ranged from 99.0%[16] to 99.9%,[14] with 81.6% (n=12,727/15,605) true negatives and 0.7% (n=111/15,605) false negatives (**Table 1**).

Among the 12,727 true negatives, 63.8% were excluded for a single reason and 36.2% for multiple reasons. Among the 111 false negatives (i.e., records incorrectly excluded by the model), all but one were excluded based on a single reason. Specifically, out of 111 false negatives, 110 (99.1%) had only one exclusion reason assigned by the model, while just one record was excluded based on multiple reasons (**Table 2**).

**Table 2:** Number of exclusion reasons among records excluded by the model.

|  | TNs | FNs | Total |
| --- | --- | --- | --- |
| **Excluded based on a single reason** | 8122 (63.8%) | 110 (99.1%) | 8,232 |
| **Excluded based on multiple reasons** | 4605 (36.2%) | 1 (0.9%) | 4,606 |
| **Total** | 12,727 | 111 | 12,838 |

FN, false negative; TN, true negative.

### Performance of AI-Assisted Screening

Across the five reviews, the model demonstrated a sensitivity of 83.2%, a specificity of 85.2%, and an accuracy of 85.1% (**Fig 2**). While the NPV was 99.1%, the model’s PPV was 19.8%.

**Fig 2:**
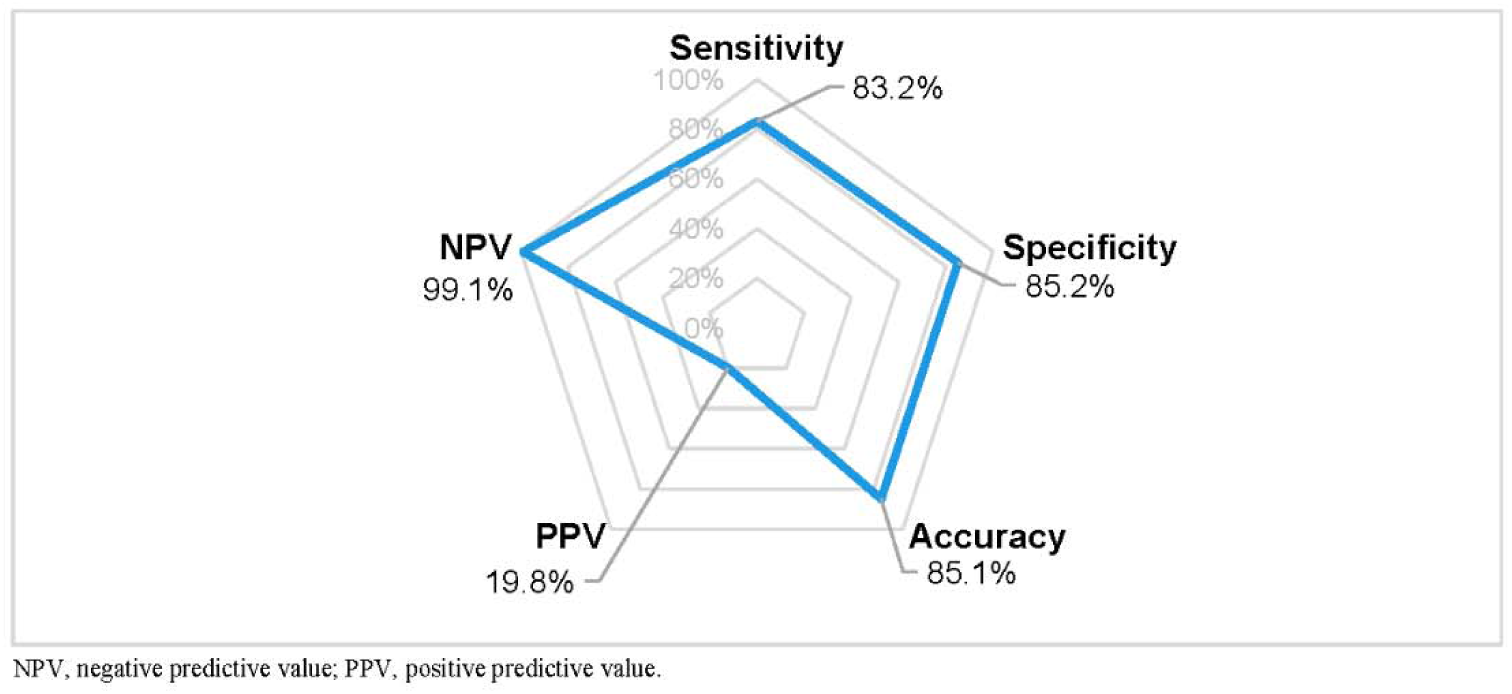
Overall performance metrics of the model for title/abstract screening

Review-level performance varied based on the nature of each systematic review, though there were no apparent trends based on the number of abstracts screened in each review (Table 1). The model achieved its highest sensitivity (97.2%) for the review by Allen et al.,[12] which focused on the humanistic burden in children with type 1 diabetes. The specificity was 75.1%, and the accuracy was 75.6%. The NPV was 99.9%, whereas the PPV for this review was one of the lowest at 7.2%. Notably, the false-positive rate was one of the highest at 24.4%, with a false-negative rate of 0.1%.

A similar pattern was observed for Bruhn et al.,[13] which assessed humanistic burden in schizophrenia; while the model achieved a sensitivity of 84.3%, specificity was 74.3%, and accuracy was 74.4%. While the NPV was 99.7%, the PPV was 4.8%. The proportion of false positives for this review was 25.3%, whereas the false-negative rate was 0.2%.

By contrast, for the review by Ejzykowicz et al.[14] on the clinical burden of renal cell carcinoma, the model reported the highest specificity at 98.4%, with a sensitivity of 94.9%, and an accuracy of 98.3%. This review reported the highest PPV observed across studies at 54.4%, and an NPV of 99.9%. The rate of false positives was 1.6%, and that of false negatives was 0.1%.

For the review by Guo et al.,[15] which focused on clinical burden related to mobility impairment, the model reported the lowest sensitivity across reviews at 76.2%, with a specificity of 88.8% and accuracy of 88.0%. The NPV was 98.3%, while the PPV was 30.7%. The false-positive rate was 10.6%, and the false-negative rate was 1.5%.

For the health state utility values review by Pourrahmat et al.,[16] the model reported a sensitivity of 93.4%, specificity of 80.6%, and accuracy of 82.0%. The NPV was 99.0%, and the PPV was 36.3%. For this review, the proportion of false positives was 17.3%, whereas the proportion of false negatives was 0.7%.

### Efficiency Comparison

On average, the model processed approximately 25 records per minute, equating to a screening time of 40 minutes per 1,000 records. In contrast, a human reviewer required approximately 16 hours, or 960 minutes, in total to screen the same number of records. This results in time savings of approximately 920 minutes per 1,000 records when the model conducts the screening. However, this estimate does not account for the additional time required for human quality control, as human review of the AI-generated output is recommended to ensure accuracy and maintain high screening standards.

## Discussion

This study evaluated the use of the GPT-4o mini model for title and abstract screening in systematic reviews, comparing its performance with traditional manual screening. The findings highlight the model’s ability to identify relevant abstracts and its adaptability to complex PICO frameworks across diverse research questions. This study contributes to the growing body of literature supporting the application of AI in SLR workflows, emphasizing its potential to enhance the accuracy, reproducibility, and scalability of evidence synthesis.

The GPT-4o mini model achieved high sensitivity and specificity in selecting studies aligned with SLR objectives and PICO criteria, indicating a strong balance between minimizing false exclusions and avoiding irrelevant inclusions. This performance suggests that AI-assisted screening can substantially reduce reviewer burden. However, it is important to note that title and abstract screening does not produce the final set of publications included in systematic reviews; all identified records for inclusion still require full-text review to confirm eligibility.

Notably, the model demonstrated higher inclusion rates in two of the five reviews evaluated, both of which evaluated humanistic burden and applied less restrictive PICO eligibility criteria during title and abstract screening. Specifically, they only applied criteria for population without specified interventions or comparators. In contrast, the other three reviews focused on interventional or clinical burden topics and used more restrictive PICO criteria, such as specific interventions and/or comparators. This distinction suggests that both the type of review and the restrictiveness of its eligibility criteria influence model behavior. Broader criteria, particularly in humanistic burden reviews, may enhance sensitivity, while more restrictive criteria may improve precision.[18] These findings underscore the importance of aligning AI screening tools with the specific methodological requirements of each review and ensuring appropriate quality control processes to maintain accuracy and consistency.

Although the overall false-negative rate was low, the model did fail to include some relevant publications. However, the vast majority of these false negatives were excluded based on a single exclusion reason, with only one instance involving multiple exclusion reasons. This pattern suggests that single-reason exclusions carry a higher risk of incorrect classifications, whereas records excluded for more than one reason are more likely to represent a correct exclusion. This finding could support a hybrid AI-human screening approach, in which human reviewers focus primarily on verifying records excluded for only one reason, while records excluded for two or more reasons could be deprioritized or accepted as final. Such an approach offers a targeted and efficient approach to quality control.

The broader landscape of AI-assisted screening highlights both efficiencies and trade-offs. For instance, Guo et al. (2024) reported a high specificity (91%) but lower sensitivity (76%) using GPT-4 for title and abstract screening,[19] highlighting a risk of missing relevant studies. In contrast, the model in the current study achieved higher sensitivity, consistent with findings from other research indicating that AI-driven screening tends to prioritize inclusivity in systematic reviews.[20–22] Hamel et al. (2020) used a machine-learning-based prioritization tool that reduced screening burden but reported a median false-negative rate of 4.6%, indicating some relevant records were missed.[20] Abogunrin et al. (2022) applied support vector machines across ten reviews, achieving a sensitivity between 90% and 100%, but with low PPVs (2-37%).[21] The authors concluded that support vector machines alone may not be sufficient. Similarly, Carey et al. (2022) used a text-mining tool and reported a sensitivity of 91% but a lower specificity of 72%.[22] The false-negative rate was 9%, suggesting some relevant records were incorrectly excluded. These findings collectively underscore the trade-off between sensitivity and specificity, a common challenge across AI screening tools.

In terms of efficiency, the model processed 1,000 titles and abstracts in 40 minutes, making it approximately 24 times faster than a human reviewer, who would require an estimated 16 hours to manually screen the same number of records. While this highlights a substantial time-saving advantage, human oversight remains essential for ensuring accuracy. A targeted hybrid approach could help balance efficiency and accuracy: following the model’s initial screening, a human reviewer could focus their review specifically on records excluded for a single reason to ensure no relevant studies are missed. In the current study, approximately 53% of records (8,232 of 15,605) fell into this category. Applying this approach, a human reviewer would need to screen only 530 records per 1,000, taking approximately 8.5 hours compared to 16 hours for full manual screening. This selective quality control approach maintains the efficiency gains of AI while minimizing the risk of false exclusions, reinforcing the value of AI as a supportive tool in the SLR process.

While AI-driven screening models, such as GPT-4o mini, can enhance efficiency, it is important to recognize that continuous refinement is necessary to optimize sensitivity and specificity. While AI can handle large-scale data processing and identify a broad set of potentially relevant studies, this increased inclusivity often comes at the cost of lower specificity, leading to the inclusion of more irrelevant records. This trade-off between sensitivity and specificity is a well-documented challenge in AI-driven systematic reviews, with previous studies highlighting that AI models effectively retrieve many relevant publications but also a substantial number of false positives.[21–23] As such, continuous refinement is necessary to improve both sensitivity and specificity. Given that GPT models are regularly updated, AI-driven screening should be viewed as a collaborative tool that complements human judgment, rather than a standalone replacement. This approach ensures that systematic reviews remain both comprehensive and efficient.

### Strengths and Limitations

This study utilizes multiple published systematic reviews as a gold standard, enabling a comprehensive evaluation of the GPT-4o mini model across diverse datasets. This approach ensures a robust comparison of AI screening results with reconciled human reviewer decisions. Additionally, the GPT-4o mini model also facilitates the processing of large volumes of literature with increased speed compared to human reviewers. This is particularly important considering the rapid growth of medical research publications. By providing empirical evidence on the model’s capabilities, this study supports the continued advancement of AI in systematic reviews and aligns with other research that underscores AI’s potential to streamline complex data processing tasks across various medical fields.[7, 8, 24, 25]

The systematic reviews selected for this evaluation were intentionally complex, focusing on specialized topics such as humanistic burden, clinical burden, and health state utility values. Given that reviews of interventional studies typically have more refined, straightforward inclusion criteria, we anticipate that the GPT-4o mini model would perform even more effectively in such contexts. The strong performance observed in the current study suggests that this approach could be particularly beneficial for systematic reviews with clearly defined criteria.

However, several limitations could be addressed to expand the scope and applicability of our findings. One notable limitation is that the model did not achieve perfect sensitivity, resulting in the exclusion of some relevant studies. Hence, relying solely on the model for screening could compromise the comprehensiveness of the systematic review, underscoring the importance of incorporating human oversight. The proposed hybrid human-AI approach can help alleviate this limitation. Another notable limitation is the model’s inconsistent specificity; although it effectively identifies relevant articles, it also tends to include a larger number of irrelevant publications. This highlights the need for human validation in subsequent screening stages. Despite these limitations, the total time required for AI-assisted screening, including human quality control, remains considerably lower than the time needed for human reviewers to conduct the entire screening process manually, which underscores the efficiency benefits of AI integration.

Finally, while the evaluation incorporated a range of datasets, the generalizability of the findings may be limited. The performance of AI systems, including GPT-4o mini, can vary significantly across diverse types of medical literature and research topics. As a result, the findings may not uniformly apply to all systematic reviews, especially those in specialized medical subfields. Expanding research to explore AI-driven approaches in additional domains will help determine whether similar performance patterns hold across various types of reviews.

### Future Directions

Future research should prioritize refining AI models to improve specificity without compromising sensitivity. The variability in specificity underscores the need for iterative model optimization using structured systematic review datasets to ensure more consistent and reliable decision-making. Additionally, exploring hybrid AI-human models that incorporate active learning frameworks could provide valuable opportunities for improvement. These models would allow continuous refinement based on human feedback, enhancing the system’s performance over time. As AI technology evolves, there is significant potential for its role in systematic reviews to expand, offering more sophisticated applications across all stages of the review. In particular, incorporating AI into data synthesis and quality assessment could significantly streamline the review process, improving both its speed and accuracy while reducing labor-intensive efforts.

## Conclusions

Our customized GPT-4o mini model for screening based on article titles and abstracts demonstrated a strong performance, effectively identifying relevant publications for systematic reviews, particularly for complex topics such as humanistic and clinical burden. The AI-driven tool identified a high number of relevant publications but showed variable specificity and included more irrelevant publications compared to traditional manual approaches. This underscores the importance of human oversight to filter out irrelevant articles in subsequent screening stages.

Furthermore, the model effectively handled systematic reviews with complex PICO frameworks, showcasing its adaptability across diverse research questions. By automating the initial, labor-intensive screening stages and maintaining high sensitivity, AI tools like GPT-4o mini can significantly streamline the review process. Ongoing improvements and customization of these tools are essential to fully realize their potential and ensure that systematic reviews remain both robust and comprehensive.

## Declarations

### Ethics approval and consent to participate

The study is solely aimed at advancing knowledge and understanding in the field of AI-based data extraction, without involving any human subjects or sensitive data. Therefore, ethical considerations and associated procedures do not apply to this research.

### Availability of data and materials

The data supporting the findings of this study are derived from publicly available sources. All data generated or analyzed as part of this study, including the prompts used for model evaluation and the corresponding model outputs, are provided within the manuscript and its supplementary files.

## Authors’ contributions

**Mir Sohail Fazeli**: Conceptualization; methodology; review and editing; supervision.

**Ellen Kasireddy**: Investigation; methodology; validation; formal analysis; writing – review and editing; project administration.

**Mir-Masoud Pourrahmat**: Project administration; review and editing.

**Cuthbert Chow**: Investigation; methodology; software; data curation; writing – review and editing.

**Jean-Paul Collet**: Investigation; methodology; review and editing; supervision.

## Acknowledgements

The authors would like to thank Michael del Aguila, Walid Shouman, Kimberly Hofer, and Jun Collet of Evidinno Outcomes Research Inc. (Vancouver, BC, Canada) for their help in validation of the results, as well as the development and submission of the manuscript.

## Supporting information

**S1 File.** Table S1 Code Snippet to Generate Title Abstract Screening Decision

**S1 File.** Table S2 PICO Criteria for Allen et al.[12].

**S1 File.** Table S3 PICO eligibility criteria for Bruhn et al.[13].

**S1 File.** Table S4 PICO Criteria for Ejzykowicz et al.[14].

**S1 File.** Table S5 PICO Criteria for Guo et al.[15].

**S1 File.** Table S6 PICO Criteria for Pourrahmat et al.[16].

## References

1. Linares-Espinós E, Hernández V, Domínguez-Escrig JL, Fernández-Pello S, Hevia V, Mayor J, et al. Methodology of a systematic review. Actas urologicas espanolas. 2018;42(8):499–506.

2. Gopalakrishnan S, Ganeshkumar P. Systematic Reviews and Meta-analysis: Understanding the Best Evidence in Primary Healthcare. Journal of family medicine and primary care. 2013;2(1):9–14.

3. Pollock A, Berge E. How to do a systematic review. International journal of stroke: official journal of the International Stroke Society. 2018;13(2):138–56.

4. Borah R, Brown AW, Capers PL, Kaiser KA. Analysis of the time and workers needed to conduct systematic reviews of medical interventions using data from the PROSPERO registry. BMJ Open. 2017;7(2):e012545.

5. van Dinter R, Tekinerdogan B, Catal C. Automation of systematic literature reviews: A systematic literature review. Information and Software Technology. 2021;136:106589.

6. Bannach-Brown A, Przybyła P, Thomas J, Rice ASC, Ananiadou S, Liao J, et al. Machine learning algorithms for systematic review: reducing workload in a preclinical review of animal studies and reducing human screening error. Systematic Reviews. 2019;8(1):23.

7. Feng Y, Liang S, Zhang Y, Chen S, Wang Q, Huang T, et al. Automated medical literature screening using artificial intelligence: a systematic review and meta-analysis. Journal of the American Medical Informatics Association: JAMIA. 2022;29(8):1425–32.

8. Khalil H, Ameen D, Zarnegar A. Tools to support the automation of systematic reviews: a scoping review. Journal of clinical epidemiology. 2022;144:22–42.

9. Clark J, Glasziou P, Del Mar C, Bannach-Brown A, Stehlik P, Scott AM. A full systematic review was completed in 2 weeks using automation tools: a case study. Journal of clinical epidemiology. 2020;121:81–90.

10. Disher T. HTA239 Streamlining Systematic Review Feasibility Assessments With Large Language Models: A Novel AI-Driven Workflow. Value in Health. 2024;27(12):S401.

11. Li M, Sun J, Tan X. Evaluating the effectiveness of large language models in abstract screening: a comparative analysis. Systematic Reviews. 2024;13(1):219.

12. Allen V, Bascle S, Cherkas A, Kasireddy E, Min R, Pushkarna D, et al. PCR86 Humanistic Burden of Informal Caregivers of Children and Young Adults With Newly Diagnosed Type 1 Diabetes (T1D): A Systematic Literature Review (SLR). Value in Health. 2022;25:S407.

13. Bruhn D, Hwang S, Howarth A, Dubé S. The burden of illness for patients with schizophrenia and primary negative symptoms: A systematic literature review. Schizophrenia research. 2022;248:341–4.

14. Ejzykowicz F, Kurt M, Dyer M, May JR, Shouman W, Kasireddy E, et al. CO33 A Systematic Literature Review (SLR) of Comparative Efficacy Measures in Randomized Controlled Trials (RCTS) of Adjuvant Treatment in Localized Renal Cell Carcinoma (RCC). Value in Health. 2022;25(12):S23.

15. Guo CC, Chiesa PA, de Moor C, Fazeli MS, Schofield T, Hofer K, et al. Digital Devices for Assessing Motor Functions in Mobility-Impaired and Healthy Populations: Systematic Literature Review. Journal of medical Internet research. 2022;24(11):e37683.

16. Pourrahmat MM, Kim A, Kansal AR, Hux M, Pushkarna D, Fazeli MS, et al. Health state utility values by cancer stage: a systematic literature review. The European journal of health economics: HEPAC: health economics in prevention and care. 2021;22(8):1275–88.

17. Higgins JP, Savović J, Page MJ, Elbers RG, Sterne JA. Assessing risk of bias in a randomized trial. Cochrane handbook for systematic reviews of interventions. 2019:205–28.

18. Fabiano N, Gupta A, Bhambra N, Luu B, Wong S, Maaz M, et al. How to optimize the systematic review process using AI tools. JCPP Advances. 2024;4(2):e12234.

19. Guo E, Gupta M, Deng J, Park YJ, Paget M, Naugler C. Automated Paper Screening for Clinical Reviews Using Large Language Models: Data Analysis Study. Journal of medical Internet research. 2024;26:e48996.

20. Hamel C, Kelly SE, Thavorn K, Rice DB, Wells GA, Hutton B. An evaluation of DistillerSR’s machine learning-based prioritization tool for title/abstract screening – impact on reviewer-relevant outcomes. BMC Medical Research Methodology. 2020;20(1):256.

21. Abogunrin S, Queiros L, Witzmann A, Sumner M, Wehler P, Baehrens D. POSA318 Automation of Title and Abstract Screening: CAN Robots Replace Humans? Value in Health. 2022;25(1):S201.

22. Carey N, Harte M, Mc Cullagh L. A text-mining tool generated title-abstract screening workload savings: performance evaluation versus single-human screening. Journal of clinical epidemiology. 2022;149:53–9.

23. Salinas MP, Sepúlveda J, Hidalgo L, Peirano D, Morel M, Uribe P, et al. A systematic review and meta-analysis of artificial intelligence versus clinicians for skin cancer diagnosis. npj Digital Medicine. 2024;7(1):125.

24. Tóth B, Berek L, Gulácsi L, Péntek M, Zrubka Z. Automation of systematic reviews of biomedical literature: a scoping review of studies indexed in PubMed. Systematic Reviews. 2024;13(1):174.

25. Santos ÁOD, da Silva ES, Couto LM, Reis GVL, Belo VS. The use of artificial intelligence for automating or semi-automating biomedical literature analyses: A scoping review. Journal of biomedical informatics. 2023;142:104389.

