## Supplementary File 1 for "Automating Screening of Titles and Abstracts in Systematic Reviews: An Assessment of GPT-4o mini"

**Supplementary Table S1: Code Snippet to Generate Title Abstract Screening Decision**

\# Title-Abstract Screening Using Large Language Models

\# Simplified pseudocode for systematic literature review screening

\# Define structured output schema for screening decisions

screening_schema = {

"papers": [{

"title": "Verbatim title of the paper",

"identifier": "Unique identifier from input",

"screening_output": {

"inclusion_reason": "Detailed reasoning about PICO criteria alignment (2-3 sentences)",

"decision": "YES or NO",

"exclusion_reasons": ["Population", "Intervention", "Comparators",

"Study Design", "Outcomes", "Duplicate Publication"]

}

}]

}

\# System prompt for LLM-based screening

BASE_PROMPT = """

You are an expert at screening medical research papers for inclusion

in a systematic literature review.

You will be given research papers with titles and abstracts. Screen each

paper for inclusion based on the provided PICO criteria, deciding whether

to include or exclude, with reasoning for the decision.

PICO Criteria: {pico_criteria}

Screening Guidelines:

- Study should satisfy ALL PICO criteria to be included

- Do not exclude solely because of mixed populations without subgroup data

in the abstract; full text may contain relevant stratified analyses

- If study design cannot be determined from title/abstract, do not use

this as grounds for exclusion

- Flag duplicate publications for exclusion

- Provide detailed reasoning for ALL decisions (include or exclude)

"""

def screen_title_abstracts(papers, pico_criteria, strictness="medium"):

"""

Screen papers using LLM with structured output parsing.

Args:

papers: List of {title, abstract, identifier} dictionaries

pico_criteria: PICO framework criteria for the review

Returns:

List of screening decisions with reasoning

"""

prompt = BASE_PROMPT.format(pico_criteria=pico_criteria)

\# Call LLM with structured output enforcement

response = llm.generate(

prompt=prompt,

context=papers,

output_schema=screening_schema

)

return response.papers

**Supplementary Table S2:** PICO Criteria for Allen et al.[12]

| **PICO item** | **Inclusion criteria** | **Exclusion criteria** |
| --- | --- | --- |
| **Population** | Informal caregivers or parents of children and young adults (ages 6 to 21), who have been newly diagnosed with T1D within approximately three months (including type 1a and type 1b) | - Formal caregivers - Studies including mixed diagnoses will be excluded, unless subgroup data is reported for T1D caregivers or care partners |
| **Interventions/ Comparators** | Any or none | N/A |
| **Outcomes** | Patient disease/symptom impact on caregiver/care partner  Outcomes regarding informal caregiver/care partner perspective, symptoms, and concepts of importance, including but not limited to:   - Family Adaptation and Cohesion Evaluation Scale (FACES) - Days absent from work (e.g., absenteeism and presenteeism) - Hospital anxiety and depression scale (HADS) - Zarit Burden Scale - CareQoL - Caregiver Reaction Assessment (CRA) - Kingston Caregiver Stress Scale (KCSS) - Caregiver burden inventory - Caregiver strain index - Burden scale for family caregivers - Caregiver burden scale - Folkman’s 4-item measure of finding positive meaning in caregiving - Health-related quality of life (HRQoL) - Beck Depression Inventory; Beck Hopelessness Scale - Chalder Fatigue Scale - Physical Fatigue subscale - 6-item Short Form Health Survey Questionnaire - 6-item Short Form Health Survey Questionnaire Mental Component - 36-item Short Form Health Survey Questionnaire Physical Component - Zung Depression Scale - State-Trait Anxiety Inventory - Utrecht Coping List Passive Approach subscale   Objective outcomes:   - Missed days at work in the past year due to caregiving responsibilities - Loss of job/employment - Lost income - Sleep quality/loss of sleep | Outcomes not directedly measuring or providing insight to caregiver/care partner burden |
| **Study design** | - Randomized controlled trials - Non-randomized clinical trials - Observational studies (including cohort studies, case-control, and cross-sectional studies) - Qualitative research (e.g., focus groups, interviews)   For library:   - Systematic literature reviews, with or without meta-analyses | - Narrative reviews - Guidelines - Case reports/series - Editorials, Notes, and Letters |

Note: Outcome-based exclusions were not permitted during title and abstract screening, following the same inclusion and exclusion criteria as the original review.

**Supplementary Table S3:** PICO eligibility criteria for Bruhn et al.[13]

| **PICO item** | **Inclusion criteria** | **Exclusion criteria** |
| --- | --- | --- |
| **Population** | Patients with schizophrenia and negative symptoms | Patients with positive symptoms of schizophrenia |
| **Interventions/ Comparators** | Any or none | N/A |
| **Outcomes** | Epidemiology outcomes   - Incidence - Prevalence - Mortality - Morbidity   Clinical outcomes, including but not limited to:   - Proportion of patients with comorbidities such as systemic disorders, hypothyroidism, diabetes, alcohol, or substance abuse - Charlson Comorbidity Index - Disability-adjusted life years (DALYs)/Years lived with disability (YLD) - Clinical Assessment Interview for Negative Symptoms (CAINS) - 4-Item Negative Symptom Assessment - PANSS (Positive and Negative Symptom Scale) - PANSS Marder Negative Symptoms Factor - Clinical burden related to polypharmacy - Schizophrenia Care and Assessment Program Health Questionnaire   Clinical burden related to non-adherence   - Medication possession ratio - Morisky Medication Adherence Scale   HRQoL outcomes, including but not limited to:   - Schizophrenic scales (e.g. Quality of Life Enjoyment and Satisfaction Questionnaire, Schizophrenia Quality of Life Scale - Generic scales (e.g. SF-36, SF-12, EQ-5D, WHOQOL, PSP) - Caregiver burden scales (e.g. Caregiver Burden Inventory, Zarit Burden Interview)   Cost outcomes, including but not limited to:   - Direct costs - Total direct medical care cost - Total costs associated with drugs and treatments and the included elements - Other costs (e.g. inpatient or outpatient care, emergency room visits, procedures, physician visits, diagnostic/screening services, rehabilitation in a facility or at home, community-based services [i.e., social supports], medical devices, aids and appliances, alternative care) - Indirect costs - Total indirect cost - Patient productivity loss - Absenteeism and presenteeism - Total out-of-pocket costs and elements included (e.g. copayments for drugs, home adaptations, aids and specialty assistive devices, personal assistance, special transportation, informal care)   Healthcare resource utilization outcomes, including but not limited to:   - Hospitalization (no. of hospitalization, length of stay, etc.) - Emergency Department visits | N/A |
| **Study design** | Observational studies   - Case-control - Prospective cohort - Retrospective cohort - Cross-sectional | - Pre-clinical studies - Case reports - Guidelines - Non-economic modelling studies - Randomized controlled trials - Non-randomized clinical trials - Single-arm trials |

Note: Outcome-based exclusions were not permitted during title and abstract screening, following the same inclusion and exclusion criteria as the original review.

**Supplementary Table S4:** PICO Criteria for Ejzykowicz et al.[14]

| **PICO item** | **Inclusion criteria** | **Exclusion criteria** |
| --- | --- | --- |
| **Population** | - Adults with localized (Stage I-III) renal cell carcinoma who have undergone partial or radical nephrectomy | Advanced or metastatic RCC (Stage IV) |
| **Intervention** | - Any treatments in the adjuvant setting, including but not limited to: - Systemic treatments - Radiotherapy - Chemoradiation | - N/A |
| **Comparator** | - Any | - N/A |
| **Outcomes** | - Overall survival (OS) ***and***   Disease-free survival (DFS), ***or*** alternatives such as relapse-free survival (RFS), ***or*** event-free survival (EFS), ***or*** progression-free survival (PFS) where definitions may overlap with that of DFS | - N/A |
| **Study design** | - Randomized controlled trials (RCTs)   For library:  Systematic reviews and meta-analyses will be included for cross-checking their included studies | - Observational studies - Narrative reviews |

Note: Outcome-based exclusions were not permitted during title and abstract screening, following the same inclusion and exclusion criteria as the original review.

**Supplementary Table S5:** PICO Criteria for Guo et al.[15]

| **PICO item** | **Inclusion criteria** | **Exclusion criteria** |
| --- | --- | --- |
| **Population** | - Healthy or unhealthy adults (>18 years) | - Children (<18 years) |
| **Interventions** | - Digital and/or wearable technologies used or being tested for assessing motor functions in healthy or unhealthy adults. - Any other intervention related to the use of digital and/or wearable devices used or being tested for assessing motor functions in healthy or unhealthy adults. | - N/A |
| **Comparators** | - Any or none | - N/A |
| **Outcomes** | - Motor function outcomes measured by digital and/or wearable technology, including by not limited to:   - Gross motor functions (e.g., gait speed, stride length, gait symmetry, bradykinesia)   - Fine motor functions (e.g., finger tapping speed, tracing accuracy)   - Oculomotor function outcomes (e.g., eye movements, pupillary reflex, blink) | - Patient-reported outcomes (PROs) (e.g., questionnaires) |
| **Study design** | - Randomized controlled trials - Non-randomized clinical trials - Observational studies   - Case-control studies   - Retrospective cohort studies   - Prospective cohort studies   - Cross-sectional studies   **For reference only (not included for data extraction):**   - Systematic literature reviews or meta-analyses - Narrative reviews | - Notes - Letters - Editorials - Comments - Case reports/ series |

Note: Outcome-based exclusions were not permitted during title and abstract screening, following the same inclusion and exclusion criteria as the original review.

**Supplementary Table S6:** PICO Criteria for Pourrahmat et al.[16]

| **PICO Item** | **Inclusion criteria** | **Exclusion criteria** |
| --- | --- | --- |
| **Population** | Patients in health states of cancer by stage  The population providing utility values could be patients with cancer (indirect measurement through quality-of-life instruments), or others such as community members (direct measurement) | - Children or adolescents - Pre- and post-progression patients* |
| **Interventions** | Any or none | N/A |
| **Comparators** | Any or none | N/A |
| **Outcomes** | Utility estimates, by cancer stage for any cancer types | N/A |
| **Study design** | Studies developing and reporting health utility values for a whole stage of cancer. Study designs may include:   - Clinical Trials - Observational studies - Surveys and data collection studies (e.g., time-trade-off studies)   **Only include for library, but not for data extraction:**   - Systematic reviews/meta-analyses of utility values in cancer state - Narrative reviews | - Case reports - Case series - Pre-clinical studies - Cost-effectiveness analyses - Cost-utility analyses - Cost-benefit analyses - Cost-minimization analyses - Cost-consequence analyses |
